# Disparities, Perceived Discrimination, and Patient-Clinician Communication in Alcohol Use Disorder Treatment: An All of Us Cohort Study

**DOI:** 10.64898/2026.02.16.26346428

**Authors:** Jungyeon Moon, J. Cesar Ignacio Espinoza, Talia Puzantian

## Abstract

**Background and Aims:** Alcohol use disorder (AUD) remains a major public health concern, with persistent disparities in access to evidence-based treatment. This study aimed to examine associations between perceived discrimination in healthcare settings (PDHS), patient-clinician communication (PCC), and receipt of treatment for AUD, and compared these with sociodemographic and insurance-related factors.

**Design:** Cross-sectional analysis using structural equation modeling (SEM), logistic and multinomial logistic regression, and machine learning approaches including SHapley Additive exPlanations (SHAP).

**Setting:** United States, using data from the National Institutes of Health All of Us Research Program.

**Participants:** A total of 5,287 adults with AUD (mean age 61 years; 57% men), including 71.6% non-Hispanic White, 12.2% Black, and 8.6% Hispanic participants. Insurance coverage included 52% government (Medicaid/Medicare), 37% private, and 21% military with 19% reporting more than one type.

**Measurements:** Primary outcomes were receipt of Food and Drug Administration-approved pharmacotherapy and/or psychotherapy for AUD, examined as binary and multinomial outcomes. The primary exposure was PDHS, measured using a 7-item scale (range 7-35), with higher scores indicating more frequent discrimination. PCC, assessed using a 2-item scale (range 2-8) with higher scores indicating poorer communication, was examined as a potential mediator. Models were adjusted for age group, sex at birth, race/ethnicity, insurance type (government, private, military), household income, and Alcohol Use Disorders Identification Test-Consumption (AUDIT-C) scores (range 0-12).

**Findings:** PDHS was associated with poorer PCC (β = 0.209, p < 0.001), although communication was not independently associated with treatment receipt. The indirect pathway from discrimination to treatment via communication was not supported. Military insurance was the strongest predictor of treatment receipt, with 6-7 times higher odds compared with other insurance types. Higher AUDIT-C scores and greater PDHS were also associated with increased likelihood of treatment. In analyses restricted to civilian participants, PDHS showed a stronger association with treatment receipt, while PCC demonstrated more modest effects. Machine learning models identified PDHS, AUDIT-C, and PCC as strong contributors, with the impact of poor communication most pronounced among individuals with lower income.

**Conclusions:** Access to treatment for alcohol use disorder is most strongly associated with insurance coverage, particularly military insurance. PDHS and PCC also contribute to treatment engagement, with differential effects across socioeconomic groups. These findings highlight the importance of addressing structural and interpersonal barriers to improve equitable access to evidence-based AUD treatment.

## INTRODUCTION

Alcohol use disorder (AUD), a chronic disease characterized by problematic alcohol consumption despite adverse social, psychological, and health consequences, not only impacts an individual’s functioning but also represents a substantial public health challenge. While overall prevalence has declined slightly in recent years, AUD remains both common and burdensome. According to the 2024 National Survey on Drug Use and Health (NSDUH), 10.3% of American adults (27.1 million individuals) met diagnostic criteria for AUD.1 Numerous significant health consequences including liver diseases, neoplasms, cardiovascular diseases, and neurologic disorders have been associated with AUD.2 AUD is considered one of the leading causes of preventable death in the United States with alcohol related deaths rising over the past decade despite effective treatments.3 Behavioral interventions, particularly cognitive-behavioral therapy, have been studied extensively and shown to reduce drinking and preventing relapse. Pharmacologic treatments such as naltrexone, acamprosate and disulfiram also have a robust evidence base supporting their role in reducing drinking and cravings and for maintaining abstinence.

However, of those who met criteria for any substance use disorder in the past year in the NSDUH, only 12.3% received any substance use treatment (eg, counseling or medication in either inpatient, outpatient, telehealth or other settings), and among those with AUD, only 2.5% received medication treatment.1 Contributing factors are multifactorial, including ambivalence about readiness to change, cost, lack of access to providers or insurance coverage, and stigma.1 Social determinants of health (SDOH) are the nonmedical factors that influence health outcomes.4 These include income, education, healthcare access (eg, insurance), housing stability, social support, among other factors, and may influence up to 60% of health outcomes.5 Discrimination based on race, socioeconomic status, or other characteristics represents a critical dimension of SDOH that contributes to health disparities and may erode trust in health care.6 A recent study examined poor patient-clinical communication (PCC) with perceived discrimination in health care settings (PDHS) and found positive association in delaying of health care for diabetes and hypertension.7 Another study examined the effects of race, ethnicity and socioeconomic status on treatment receipt for AUD and found disparities contributing to access inequity.8

In this study, we examine potential disparities including SODH, poor communication and perceived discrimination to identify their role in receipt of and perseverance in treatment of AUD. Launched in 2015, All of Us is a federal initiative to advance precision and health equity through the collection of genetic, health, and demographic data from over 1 million American volunteers including those historically underrepresented in biomedical research.9 We hypothesize that the diversity of participants along with the breadth of available data would lend to a compelling examination of connections between health disparities and perceived discrimination. Understanding potential disparities and discrimination can better inform healthcare policies to address existing barriers to treatment and thereby improve patient outcomes.

## METHODS

### Study design and data source

We conducted a cross-sectional analysis utilizing data from the NIH All of Us Research Program.^9^ The analytic sample included adults aged ≥18 years with a new diagnosis of alcohol use disorder (AUD), identified using International Classification of Diseases, Tenth Revision (ICD-10) codes (F10.x).^8^ This study used de-identified data from the All of Us Research Program and did not involve human subjects as defined by federal regulations; therefore, Institutional Review Board approval was not required. All participants provided informed consent at enrollment in the All of Us Research Program.

### Outcome variables

AUD treatment was defined as receipt of FDA-approved pharmacotherapy (naltrexone, acamprosate, or disulfiram) and/or psychotherapy for AUD.^10^ Treatment was coded both as a binary indicator (any treatment vs none) and as multinomial outcomes (medication only, psychotherapy only, both).

### Exposure

Perceived discrimination in health care settings (PDHS) was assessed using a 7-item scale derived from the Social Determinants of Health survey. The items captured the frequency of unfair treatment experiences in healthcare setting, with higher scores indicating greater perceived discrimination (range 7-35).^6,7,9,11^ Detailed item wording and scoring procedures are provided in the Supplementary Methods.

### Mediator

Patient-clinician communication (PCC) was assessed using two survey items from the Health Care Access and Utilization survey, measuring the clarity of provider communication and patient involvement in decision-making (range 2-8).^7,9,11^ Items were reverse coded so that higher scores represent poorer communication. Additional details are provided in the Supplementary Methods.

### Covariates

Models were adjusted for age group (45-64, ≥65 years), sex at birth, race/ethnicity (Black, Hispanic, other), insurance type (government, private, military, other), household income (<$75,000 vs higher), and AUD severity using the Alcohol Use Disorders Identification Test - Consumption (AUDIT-C) (range 0-12).^12^ Detailed AUDIT-C scoring procedures are described in the Supplementary Methods.

### Statistical Analysis

We combined regression-based and machine learning approaches. Structural Equation Modeling (SEM) was used to test the hypothesized mediation pathway (PDHS → PCC → treatment), with moderators (age, race/ethnicity, and insurance). Logistic and multinomial logistic regression assessed direct associations and predictors of specific treatment types. Interaction terms (PDHS×age, PDHS×race, PDHS×insurance) were tested to evaluate effect modification. Finally, random forest (rf) models with Shapley Additive exPlanations (SHAP) values were applied to explore nonlinear relationships among the tested predictors. Analyses were conducted in Python using statsmodels for regression, semopy for SEM, and scikit-learn and shap for machine learning (ML). Statistical significance was defined as p < 0.05.

## RESULTS

We identified 5,287 individuals with AUD and relevant survey and sociodemographic information (mean age 61 years, 57 % men). Approximately 12.2 % self-identified as Black, 8.6 % as Hispanic, 71.6 % as non-Hispanic White. Insurance coverage was diverse, 52 % government (Medicaid, Medicare), 36.6 % private, and 21 % military (Military, VA) with 19% reporting more than one type.

In SEMs (Figure 1 path a1, Table 1), higher PDHS scores were significantly associated with poorer PCC (β ≈ 0.209, p < .001). However, PCC was not significantly (Figure 1, path b) associated with treatment receipt, and the hypothesized indirect PDHS → PCC → Treatment pathway was not supported (Figure 1 path c, Table 1). A direct effect of PDHS on treatment remained significant, suggesting that discrimination influences treatment independently of communication.

**TABLE 1.** Coefficients from the structural equation model.

|  | Outcome |  |  |  | Treatment Engagement |  |
| --- | --- | --- | --- | --- | --- | --- |
|  |  | PCC |  |  |  |  |
| Mediator | label | Coefficient (95% CI) | P value | label | Coefficient (95% CI) | P value |
| PCC | NA | NA | NA | b | -0.010 (-0.023, -0.003) | 0.132 |
| Predictor |  |  |  |  |  |  |
| PDHS | a1 | 0.025 (0.127, -0.283) | 0.000 | c | 0.031 (0.018, -0.044) | 0.000 |
| > 64 yo | a4 | -0.209 (-0.291, -0.128) | 0.000 |  | -- | -- |
| Black | a2 | -0.130 (-0.214, -0.047) | 0.002 | Not shown | -0.065 (-0.106, -0.025) | 0.000 |
| Private Ins. | a3 | -- | -- | Not shown | -0.080 (-0.111, -0.050) | 0.000 |
| Government Ins. | a3 | -- | -- | Not shown | -0.084 (-0.112, -0.055) | 0.000 |
| Military Ins. | a3 | -- | -- | Not shown | -0.368 (0.335, 0.401) | 0.000 |
| > 75,000 / yr | Not shown | -- | -- | Not shown | -0.066 (-0.097, 0.035) | 0.000 |
| PDHs Interactions |  |  |  |  |  |  |
| 45 - 64 yo | a4 | 0.072 (0.001, 0.143) | 0.046 |  | -- | -- |
| Other Ins. | a7 | -0.205 (-0.377, -0.033) | 0.019 |  | -- | -- |

Note: Path labels correspond to Figure 1. Only statistically significant coefficients are shown, except for the PDHS → treatment path, which is included because it represents the primary hypothesized association. Coefficients are presented with 95% confidence intervals.

**FIGURE 1.**
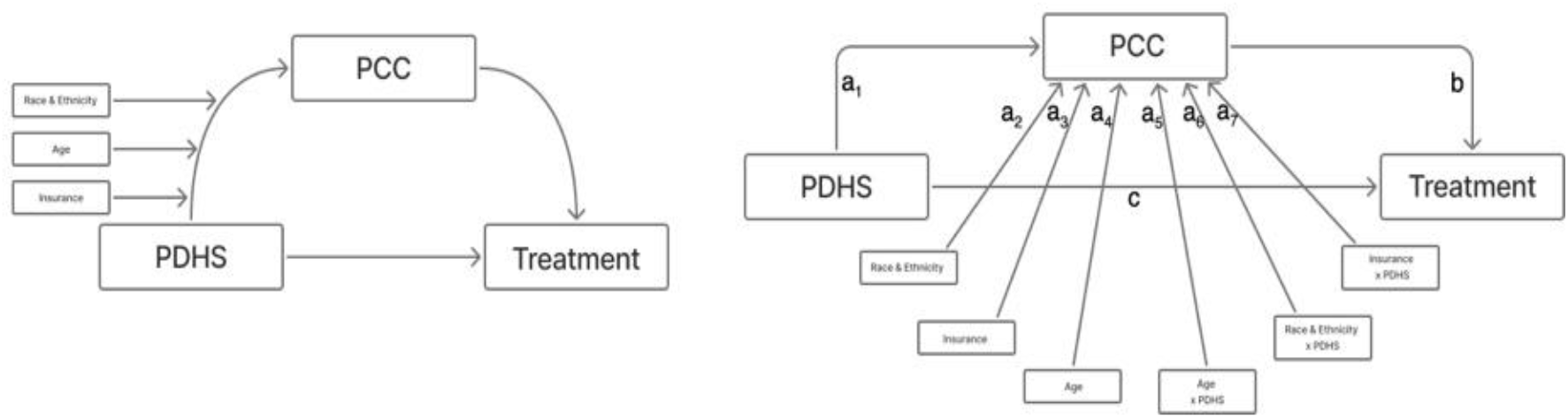
Conceptual and statistical models of the association between perceived discrimination, patient-clinician communication, and treatment receipt. The left panel illustrates the conceptual model in which patient–clinician communication (PCC) mediates the association between perceived discrimination and treatment receipt, with race/ethnicity, age, and insurance status included as covariates. The right panel presents the corresponding statistical model, with path coefficients displayed for each estimated association.

Since our initial mediation hypothesis was not supported, we tested direct effects independent of moderation and interactions on treatment. Multiclass logistic regression results (Figure 2) showed that military insurance was the strongest predictor of treatment receipt, with odds ratios ranging from approximately 6 to 7. While these findings largely align with previous reports, we also found that higher PDHS and AUDIT-C scores were associated with higher odds of treatment.

**FIGURE 2.**
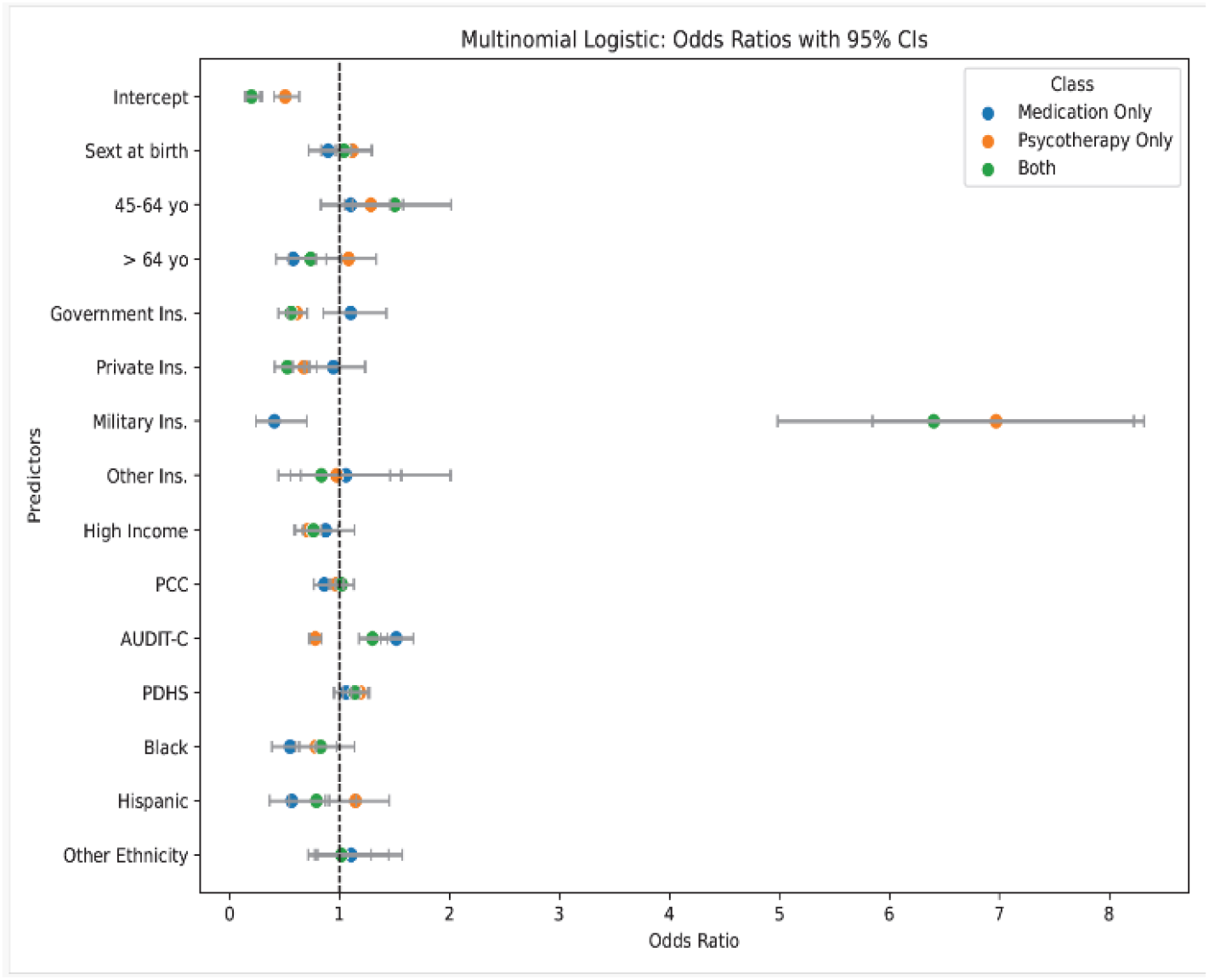
Direct effects from multivariable logistic regression models. Odds ratios (ORs) represent the associations between individual predictors and receipt of treatment. Baseline categories were female sex, age 18-44 years, no insurance, low income (<$75,000/year), and White race/ethnicity. Patient– clinician communication (PCC), Alcohol Use Disorders Identification Test-Consumption (AUDIT-C), and perceived discrimination in health care settings (PDHS) were standardized (z-scores) to facilitate comparison of effect sizes across predictors. Odds ratios greater than 1 indicate higher likelihood of receiving treatment relative to the reference group, whereas values less than 1 indicate lower likelihood. Notably, military insurance was associated with approximately six-to seven-fold higher odds of treatment receipt.

Since we were particularly interested in the effects of PDHS and PCC on treatment engagement we examined their interactions with military insurance and assessed direct effects after stratifying our cohort to include only participants with non-military insurance (Figure 3). Because the multinomial outcome became imbalanced after stratification, treatment was recategorized as a binary outcome (any treatment vs no treatment). In these analyses, military insurance remained a strong predictor in the full sample, while interaction terms (military x PDHS, military x PCC) were not significant. In civilian-only analyses, PDHS showed the strongest association with treatment receipt, and PCC contributed a moderate effect, suggesting psychosocial factors play a more prominent role outside military health systems. Finally, to better understand the characteristics of individuals for whom PDHS and PCC seem to impact treatment, we trained a random forest classifier with modest discrimination (AUC = 0.61). Feature importance analysis (Figure 4) confirmed PDHS, AUDIT-C, and PCC as strong predictors. SHAP dependence plots suggested that the effects of PCC were concentrated among low-income individuals (Figure 4).

**FIGURE 3.**
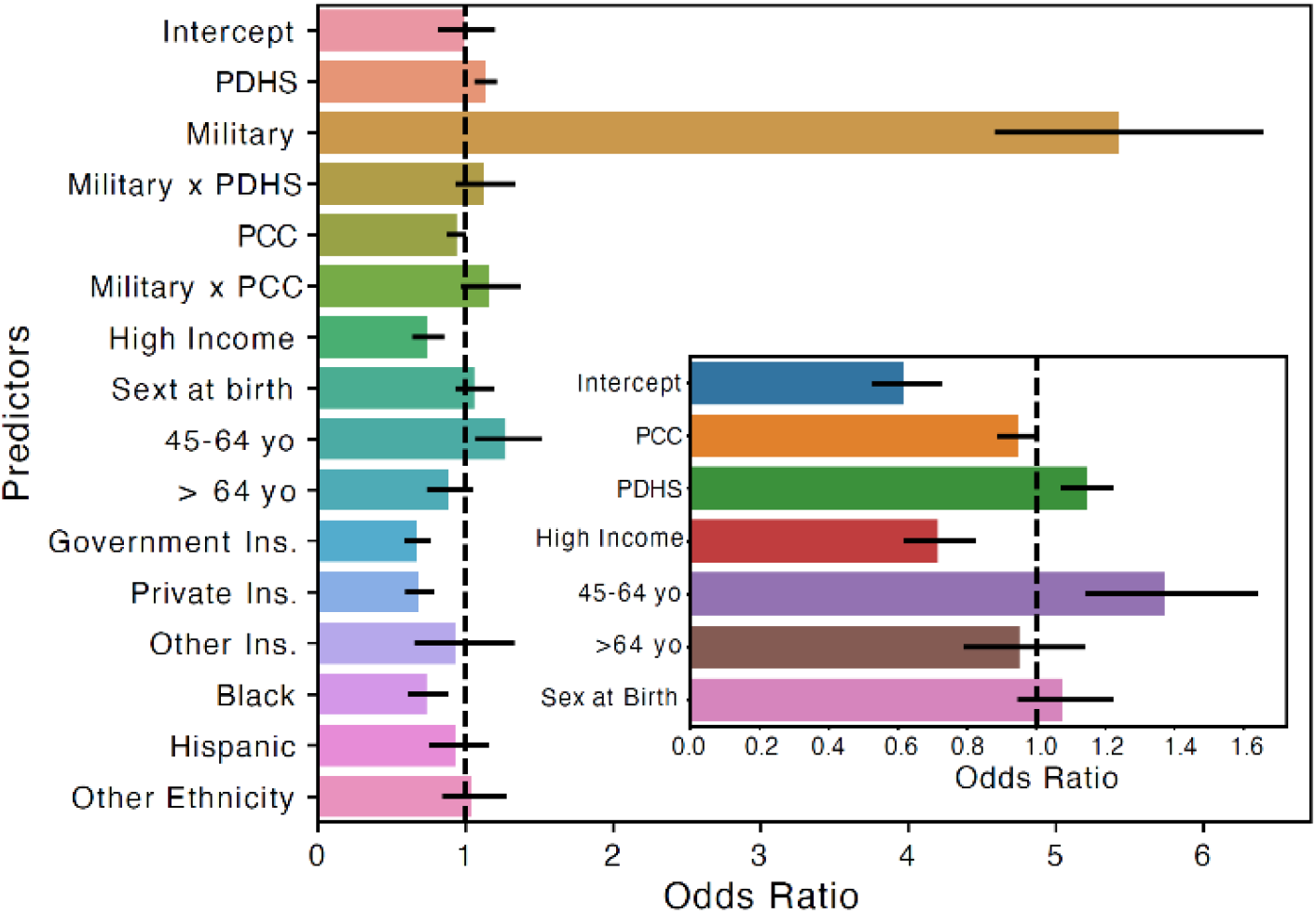
Psychosocial predictors of treatment receipt among military and civilian populations. The main panel presents results from the logistic regression model including all participants, demonstrating that military insurance is the strongest predictor of treatment receipt, with odds substantially higher than those associated with any other factor. Interaction terms between military status and perceived discrimination (PDHS) or patient–clinician communication (PCC) were centered near unity, indicating no evidence that the effects of PDHS or PCC differ by military status. The subpanel displays results restricted to civilian participants. In this model, psychosocial factors emerged as meaningful predictors of treatment receipt, with PDHS showing the strongest association and PCC contributing a moderate but consistent effect. These findings indicate that although military insurance strongly shapes access to treatment, psychosocial factors such as perceived discrimination and communication play a more prominent role among civilians.

**FIGURE 4.**
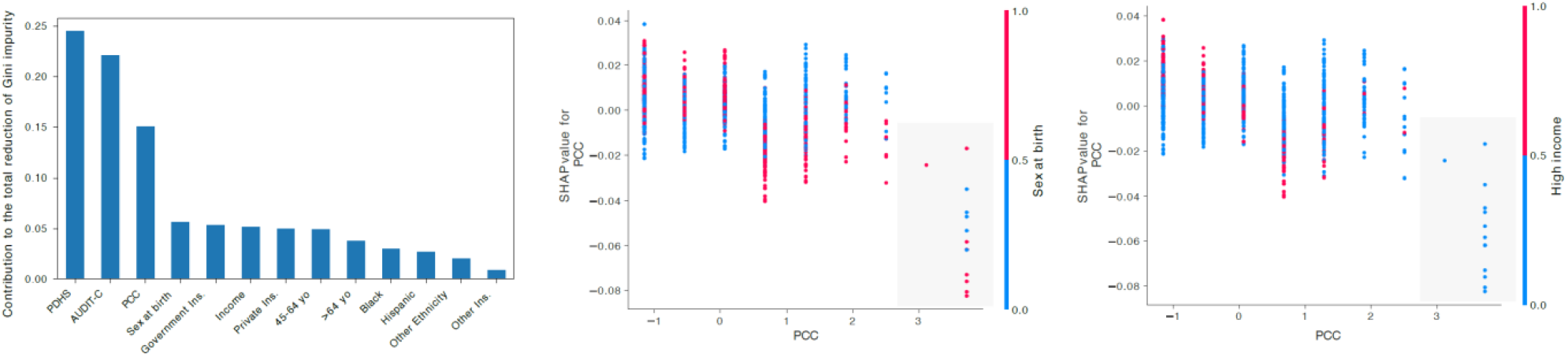
Random forest feature importance and SHAP value interactions. The random forest model was trained excluding military insurance to highlight other predictors. Feature importance analysis identified PCC, AUDIT-C, and PDHS as the most influential predictors. SHAP plots showed that poorer PCC (higher values on the x-axis) was a more effective classifier among low-income participants. Notably, the gray box in the rightmost SHAP plot contains only one class (blue dots), indicating that the interaction between PCC and income is uniquely concentrated in this group, a pattern not observed in the other plot, which is included for comparison.

## DISCUSSION

In this cross-sectional analysis of participants with AUD in the All of Us Research Program, military insurance was the strongest predictor of treatment receipt. This pattern is consistent with earlier studies showing that structured and coordinated health systems, such as the military, provide more reliable access to care. ^8,11,14,15^ This finding is also consistent with prior analyses within the All of Us cohort, which reported that approximately 70% of individuals with AUD received no treatment overall, and that military and VA coverage were associated with higher use of psychotherapy and combination treatment compared with private insurance.^8^ In contrast, racial and ethnic disparities in medication receipt were observed primarily within civilian insurance groups, emphasizing the role of structured health systems in mitigating access gaps. Together, these findings highlight the potential benefits of coordinated health systems in reducing treatment gaps for AUD.^14^ Evidence from population-based surveys further supports this pattern. Using NSDUH data, Acevedo and colleagues reported persistently low treatment utilization for AUD nationwide and considerable disparities by insurance status, particularly among Medicaid enrollees and uninsured individuals.^16^ These findings suggest that insurance structure, rather than race or ethnicity alone, is an important determinant of treatment access across settings.

Among civilians in our study, psychosocial and socioeconomic factors appeared to be more influential. Higher perceived discrimination was associated with greater treatment receipt. PCC showed modest associations overall but was most relevant among lower-income participants, indicating that communication barriers may weigh more heavily on disadvantaged groups. Higher income was unexpectedly associated with lower treatment initiation, which may reflect a combination of factors, including greater ability to self-manage or avoid formal treatment, differences in perceived need for care, or stigma that discourages treatment-seeking among more socioeconomically advantaged groups.

These findings highlight the complex and sometimes counterintuitive ways in which socioeconomic factors intersect with addiction care. These results extend prior literature on discrimination and health care utilization.^17,18^ While discrimination can occur at interpersonal, institutional, and structural levels,^19,20^ most prior studies have focused on interpersonal experiences.^21^ By applying the PDHS scale, we characterize institutional aspects of patient experiences. Previous studies show that discrimination can undermine care by delaying access, lowering satisfaction, and reducing preventive service use.^22,23^ Among people with substance use disorders, discrimination based on drug use is particularly salient, especially for Black and Hispanic populations, where it has been associated with worse mental health and more chronic disease burden.^24–26^ However, prior studies among people who use drugs suggest that although perceived discrimination is common and associated with poorer mental and physical health, only a minority identify discrimination as a direct barrier to accessing medical care, supporting the interpretation that discrimination may alter pathways to care rather than uniformly preventing treatment. Stigma is increasingly recognized as a driver of both reduced care-seeking and increased surveillance in health systems.^27,28^ One plausible mechanism is that stigma-related pathways may increase surveillance and monitoring, leading to greater exposure to treatment through more frequent interactions with healthcare and social service systems, including mandated or emergency care. This suggests that perceived discrimination may both weaken trust in health care and increase contact with health systems, particularly among populations with greater system involvement.

Previous research has shown that poor PCC mediates discrimination-related delays in chronic disease populations.^7^ Consistent with this literature, we found that PCC was most consequential for socioeconomically disadvantaged participants. We extend this work to AUD, demonstrating that interventions to improve communication quality and shared decision-making may represent a modifiable target for reducing disparities in treatment engagement. Emerging evidence suggests that digital and web-based interventions can enhance communication, support continuity of care, and reduce barriers to treatment, particularly for disadvantaged populations.^29–32^ Low-income groups often encounter additional challenges, including limited English proficiency, low health literacy, and unmet social needs.^33–35^ These factors may exacerbate health disparities, especially for individuals with multimorbidity, who face complex care demands and heavier self-management burdens.^36–37^ In contrast, high quality PCC has been associated with improved adherence, self-management, and reduced stigma.^38–40^Alongside stigma-sensitive approaches^28^ and policies that expand insurance coverage, these strategies could help reduce disparities in AUD treatment receipt. Our findings demonstrate how structural, social, and interpersonal factors influence engagement with evidence-based treatment for AUD. Insurance type represents a dominant structural determinant, while discrimination and communication emerge as relevant social and interpersonal influences, particularly outside of military health systems. Stigma may not only function as a barrier to care but also as a mechanism that increases contact with healthcare systems, highlighting the dual role of stigma in addiction treatment. The observation that communication barriers are most pronounced among low-income individuals suggests a modifiable point for intervention. Efforts to improve provider communication and shared decision-making may be especially impactful in reducing disparities for socioeconomically disadvantaged patients with AUD.

This study has several limitations. First, the cross-sectional design limits causal inference, and observed associations may be subject to unmeasured confounding. Future longitudinal analyses are needed to clarify the timing of treatment initiation and persistence. Second, PDHS and PCC were self-reported, which may introduce recall or reporting bias. In addition, the discrimination measure was based on the Everyday Discrimination Scale as implemented in the All of Us Social Determinants of Health survey, which may not capture all relevant dimensions of perceived discrimination and has recognized conceptual limitations.^20^ Third, participation in the All of Us Research Program is voluntary, and although the cohort is diverse and intentionally oversamples groups historically underrepresented in biomedical research, participants differ in several characteristics from the general U.S. population, which may limit the generalizability of prevalence estimates.^9^

In conclusion, this study demonstrates that access to AUD treatment is influenced by structural coverage, socioeconomic context and interpersonal communication. Military insurance reflects a structural model of access, whereas in civilian populations discrimination and communication appear more prominent. These findings highlight the complexity of these mechanisms and suggest application of both systemic and modifiable targets for intervention, especially for low-income individuals.

## Data Availability

The data used in this study are available from the All of Us Research Program. Access to the data requires registration and approval through the All of Us Researcher Workbench.

https://www.researchallofus.org/

## AUTHOR CONTRIBUTIONS

Jungyeon Moon: Conceptualization (lead); data curation (lead); cohort construction (lead); methodology (lead); project administration (lead); supervision (lead); writing - original draft (lead); writing - review & editing (lead).

J. Cesar Ignacio Espinoza: Formal analysis (lead); methodology (supporting); data curation (supporting); visualization (lead); writing - original draft (supporting); writing - review & editing (equal).

Talia Puzantian: Conceptualization (supporting); methodology (supporting); data curation (supporting); supervision (supporting); writing - original draft (supporting); writing - review & editing (equal).

## PRIMARY FUNDING

None

## DATA AVAILABILITY STATEMENT

The data used in this study are available through the All of Us Research Program Controlled Tier (version 8) and can be accessed by authorized researchers via the All of Us Researcher Workbench.

## ACKNOWLEDGMENTS

The authors gratefully acknowledge the participants of the All of Us Research Program for their contributions, without whom this research would not have been possible. We also thank the National Institutes of Health’s All of Us Research Program for making available the participant data examined in this study.

